# Rapid resistance detection is reliable for prompt adaptation of isoniazid resistant tuberculosis management

**DOI:** 10.1101/2023.12.15.23299856

**Authors:** Olivier Bahuaud, Charlotte Genestet, Elisabeth Hodille, Maxime Vallée, Quentin Testard, Caroline Tataï, Julien Saison, Jean-Philippe Rasigade, Gérard Lina, Florence Ader, Oana Dumitrescu, the Lyon TB study group

## Abstract

**Objectives:** Appropriate tuberculosis (TB) management requires anti-TB drugs resistance detection. We assessed the performance of rapid resistance detection assays and their impact on treatment adaptation, focusing on isoniazid resistant (HR) TB.

**Methods:** From 2016 to 2022, all TB cases enrolled in 3 hospitals were reviewed for phenotypic drug susceptibility testing (p-DST) and genotypic DST (g-DST) performed by rapid molecular testing, and next generation sequencing (NGS). Clinical characteristics, treatment and outcome were collected for HR-TB patients. The concordance between g-DST and p-DST results, and delay between treatment initiation and results of g-DST and p-DST were respectively recorded to assess the contribution of DST results on HR-TB management.

**Results:** Among 654 TB cases enrolled, 29 were HR-TB. Concordance between g-DST by rapid molecular methods and p-DST was 76.9%, whilst concordance between NGS-based g-DST and p-DST was 98.7%. Rapid resistance detection significantly fastened HR-TB treatment adaptation (median delay between g-DST results and treatment modification was 6 days). It consisted in fluoroquinolone implementation for 17/23 patients; outcome was favourable except for 2 patients who died before DST reporting.

**Conclusion:** Rapid resistance detection fastened treatment adaptation. Also, NGS-based g-DST showed almost perfect concordance with p-DST, thus providing rapid and safe culture-free DST alternative.

## Introduction

Tuberculosis (TB), caused by *Mycobacterium tuberculosis* (Mtb), is the leading cause of death from a single infectious agent worldwide ^1^. Effective TB control is threatened by the global emergence and spread of anti-TB drug resistant Mtb, which hampers antibiotics efficacy and therefore requires treatment adaptation to avoid treatment failure or relapse. Mtb drug resistance may present either as resistance to rifampicin (R) or isoniazid (H) alone, or as multi-drug resistant (MDR) TB (meaning that Mtb is resistant to both major first-line anti-TB drugs R and H) ^1^.

The resistance to H, alone or combined with other drugs, is now the most common type of resistance to anti-TB drugs; the frequency of H-resistant R-susceptible TB (HR-TB) is greater than the one of R-resistant TB, and estimated at 7.4% of new and 11.4% of previously treated TB cases ^2^. Evidence indicates increased rates of poor treatment outcomes, such as treatment failure, relapse, and progression to MDR-TB, when patients with HR-TB are treated with the standard anti-TB regimen ^3^.

To enable a prompt treatment adaptation, the World Health Organization (WHO) recommends an early detection of drug resistant TB through the use of a rapid diagnostic test ^4^. Accurate rapid detection of H resistance is challenging, as H resistance is linked to multiple genetic variations in different loci, resulting in technical difficulties, which explains the heterogeneous performance of rapid H resistance detection tests ^5^. Depending on the series and the genetic particularities of Mtb isolates tested, 30% to 70% of H resistant strains remain undetected ^6^.

Though time-consuming and requiring level 3 biological safety laboratories (BSL3), the standard reference for H susceptibility detection still relays on phenotypic culture-based drug susceptibility detection (p-DST). Testing using different critical concentrations offers the possibility to distinguish between low- and high-level of H resistance and may guide the decision for treatment regimen adaptations. *In vitro* data suggest that high-dose H may be effective for the treatment of patients having low-level of HR-TB ^7^, while high-level of HR-TB requires the replacement of H by a fluoroquinolone (FQ), mainly levofloxacin (LFX) ^3^. As recommended by the WHO, it is essential that the resistance to R is excluded and, when possible, the resistance to FQ should also be excluded, prior to the implementation of LFX containing regimen, to help avert the acquisition of additional drug resistance ^4^.

Interestingly, recent progress has been done in the field of resistance detection based on next generation sequencing (NGS) to perform targeted or whole genome sequencing (WGS). The Deeplex Myc-TB® (GenoScreen, Lille, France) NGS-based tool provides a rapid identification of a large set of mutations in different resistance associated loci, up to 13 antibiotics, including all first-line anti-TB drugs and some second-line molecules such as FQ, linezolid, bedaquilin and injectable drugs, with performances equalizing those of p-DST ^8^. Likewise, WGS allows exhaustive drug resistance detection by using a mutation database validated by the WHO^9^. These NGS based diagnosis tests not only enable accurate detection of HR-TB, but may also provide information on pyrazinamide (Z), ethambutol (E) and FQ susceptibility, enabling a secure usage of LFX as recommended by the WHO guidelines ^10^.

France is a high-income country with low prevalence of TB cases (less than 10/100,000 inhabitants); in a previous retrospective study from 2016 and 2017, only 26% of patients diagnosed with TB had rapid molecular detection of H resistance, which correlated with early adaptation of HR-TB treatment ^11^. Thus, the first objective of the present study was to explore the impact of rapid resistance detection tests, including NGS-based approach on the management of the patients and on the TB disease outcome, with a focus on HR-TB cases. The second objectives were to assess the performances of H resistance diagnosis tests and the compliance of HR-TB treatment regimens with international guidelines.

## Methods

### Data collection

This is a retrospective study conducted in one tertiary-care (Hospices civils de Lyon, France, hosting 5362 hospitalisation beds) and two secondary-care hospitals (Bourg-en-Bresse and Valence, France, hosting respectively 860 and 800 hospitalisation beds). Were included, between November 2016 and July 2022, all patients diagnosed with TB and having a positive Mtb culture, at the central laboratory equipped with BSL3 and NGS facilities (Institut des Agents Infectieux, Lyon, France). For all Mtb clinical isolates, WGS analysis was performed in routine practice as part of the laboratory diagnosis. For patients with HR-TB, demographic data (age, sex, continent of birth), clinical presentation (symptoms, comorbidities, pulmonary, extra-pulmonary TB), anti-TB treatment, outcome and microbiological data (sputum smear results, time to positivity, p-DST and genotypic DST (g-DST), lineage) were collected. All the data were implemented in a database, in accordance with the decision 20-216 of the ethics committee of the Lyon University Hospital and the French legislation in place at the time of the study (Reference methodology MR-004). Relevant approval regarding access to patient-identifiable information are granted by the French data protection agency (*Commission Nationale de l’Informatique et des Libertés*, CNIL).

### Mtb culture process

Mtb clinical isolates culture and p-DST were performed as previously described ^12^. G-DST by rapid molecular methods were performed on cultured Mtb on a routine basis as previously described, using the line probe assay GenoType MTBDRplus v2.0 test (Hain Lifescience, Nehren, Germany) from November 2016 to August 2020 ^12^ and the MDR/MTB ELITe MGB® Kit (ELITechGroup SpA, Torino, Italy) on the ELITe InGenius® platform (ELITechGroup SpA) ^13^ from September 2020 to July 2022. When clinically suspected, the R resistance was also performed using Xpert MTB/RIF Ultra (Cepheid, Sunnyvale, CA, USA) on the primary sample as recommended ^14^. Finally, all R and H resistant isolates were retrospectively analysed using Deeplex Myc-TB® (GenoScreen, Lille, France) according to the manufacturer’s instructions ^15^.

### Whole-genome sequencing and bioinformatic analysis

Genomic DNA extraction, library preparation, short-read WGS using Illumina technology (San Diego, CA, USA) and bioinformatic analyses were performed as previously described ^16^. The reference genome coverage breadth was at least 92% with a mean depth of coverage of at least 30x. Variant calling with MuTect2 in microbial mode was performed. Variant calls were processed using the open access SNP-IT tool (https://github.com/samlipworth/snpit) to identify Mtb complex lineage based on WGS SNP calling ^16^. Mykrobe ^17^ command-line version is included in the pipeline to detect antibiotic resistance. Moreover, the WHO catalogue of mutations in Mtb complex and their association with drug resistance was also used as a database to annotate variants and to detect antibiotic resistance ^9^.

### Statistical analysis

Data were expressed as count (percentage, %) for dichotomous variables and as median (interquartile range [IQR]) for continuous values. The number of missing values was excluded from the denominator. For dichotomous variables, χ2 test was used and for continuous values, the paired t-test was used to compare groups. Statistical analyses were performed using GraphPad Prism® for Windows version 5.02 (GraphPad Software, La Jolla, CA, USA). A p-value < 0.05 was considered significant.

## Results

### Microbiological characterisation of Mtb isolates

Among the 654 patients with TB diagnosed at the Institut des Agents Infectieux of Lyon University Hospital from November 2016 to July 2022, 29 (4.4%) had HR-TB, 6 (0.9%) had R resistant TB and 9 (1.4%) had MDR-TB, according to p-DST (Figure S1A and C, Table S1). HR-TB was found in all lineages of Mtb *stricto sensu* (Figure S1B). The WGS revealed that the prevalence of FQ resistance was very low, only 3 cases were detected (0.46%); 1 strain was susceptible to first-line anti-TB drugs, 1 was an HR-TB strain, and 1 was an MDR-TB strain (Figure S1D and Table S1).

A total of 39 patients presented at least an H resistance, the most common mutations were the S315T on the *katG* gene, which is associated with high-level resistance, and the C-15T on the *inhA* promoter, which is associated with low-level resistance. Herein, rapid molecular methods failed to accurately identify 5 mutations associated with H resistance, concerning 9/39 (23%) TB patients with H resistance. Three mutations on the *katG* gene, that are associated with high-level resistance, were not detected, or gave an uninterpretable result. Moreover, 2 mutations on the *inhA* gene associated with the C-15T mutation on the *inhA* promoter were considered as low-level resistance mutations instead of high-level resistance (Table 1). Regarding NGS-based methods, the Deeplex Myc-TB method just failed to identify a single nucleotide deletion at the end of the coding region of the *katG* gene, at amino acid 975. WGS correctly identified all H-resistant isolates; overall the sensitivity of NGS-based methods for H resistance detection was 98.7%.

**Table 1:**
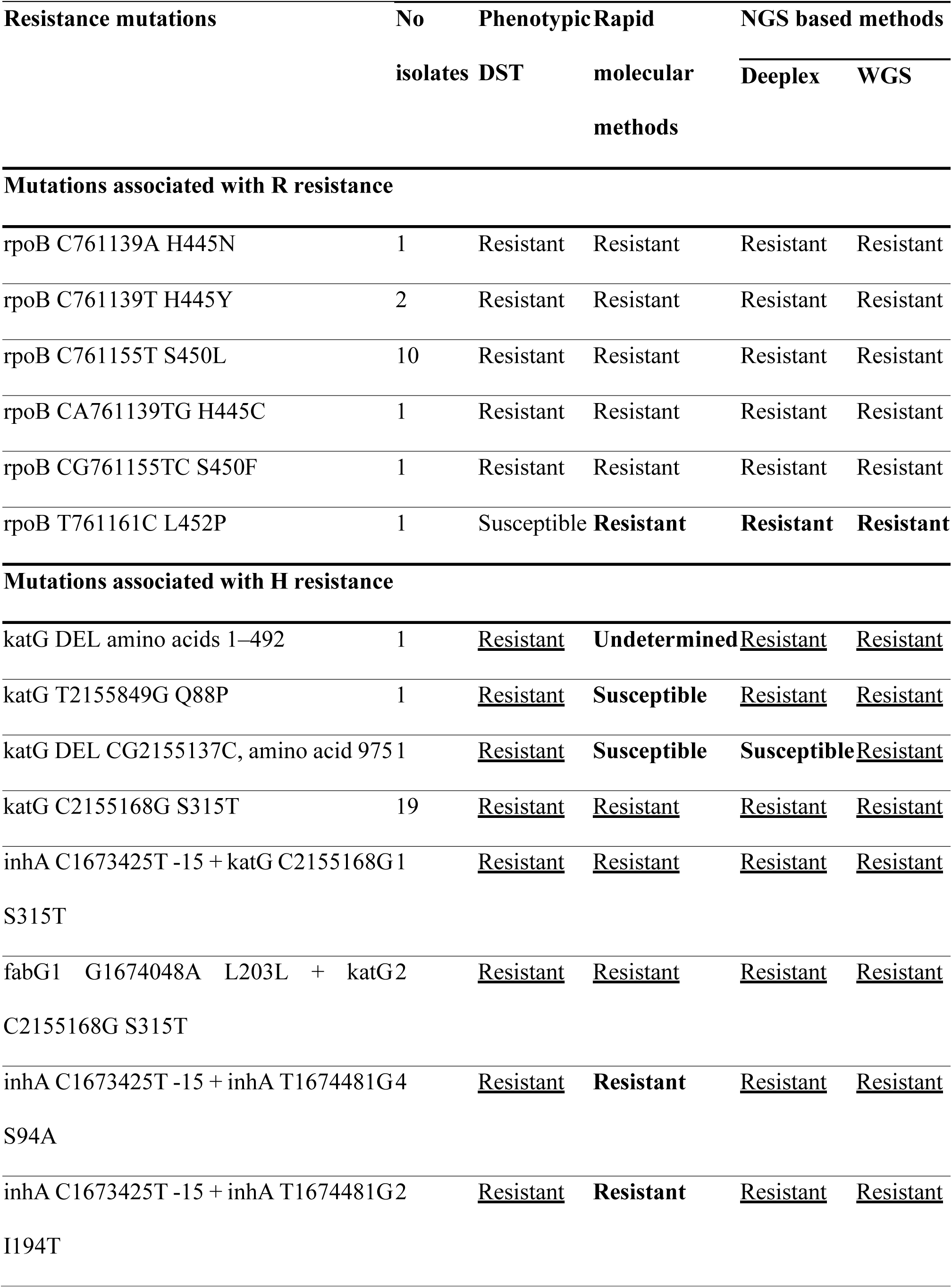
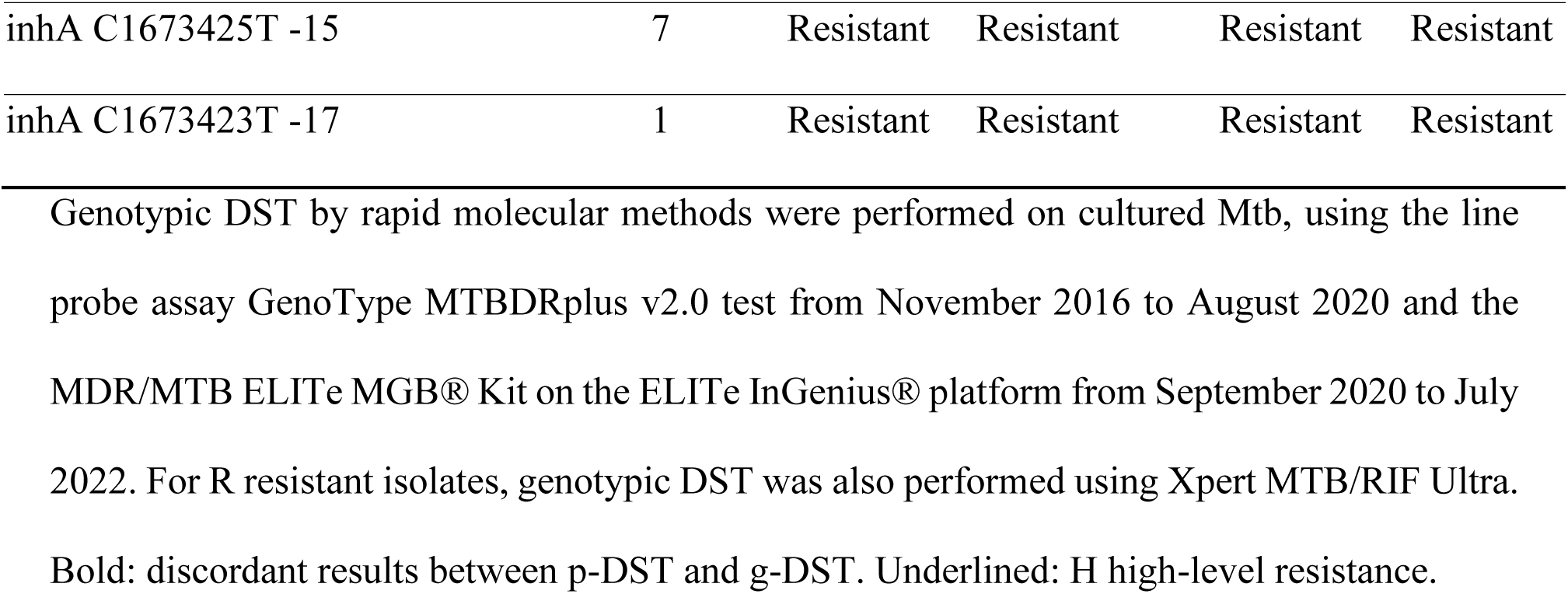
R and H resistance detection by phenotypic and genotypic methods.

Regarding R resistance, among the 6 mutations detected, the most common was the S450L mutation on the *rpoB* gene. All mutations associated with R resistance were detected by g-DST, both by rapid molecular methods and by NGS-based methods. The use of these methods even allowed to identify a low-level R resistance mutation, the L452P mutation on the *rpoB* gene, which is not always detected by p-DST (Table 1).

### Epidemiological and clinical characteristics of HR-TB patients

Among the 29 patients with HR-TB, the clinical data and information on treatment and outcome were available for 23 patients (Table S2); these data were missing for 6 patients who had been transferred to distant care centres upon TB management, and were therefore excluded from the analysis.

The median [IQR] age at diagnosis was 37 [21-53.5] years old, there were 16 (69.6%) male patients, and 19 (82.6%) were not born in France, coming from various countries within Africa, Asia, and Europe (Table 2). Five patients (21.7%) had comorbidities at TB diagnosis; 3 patients (13.0%) presented diabetes mellitus, and 2 of them (8.7%) also presented underlying chronic pulmonary conditions. Two patients (8.7%) had chronic viral hepatitis B and C, respectively, and a single patient (4.3%) was recently diagnosed with Human Immunodeficiency Virus (HIV) infection.

**Table 2:**
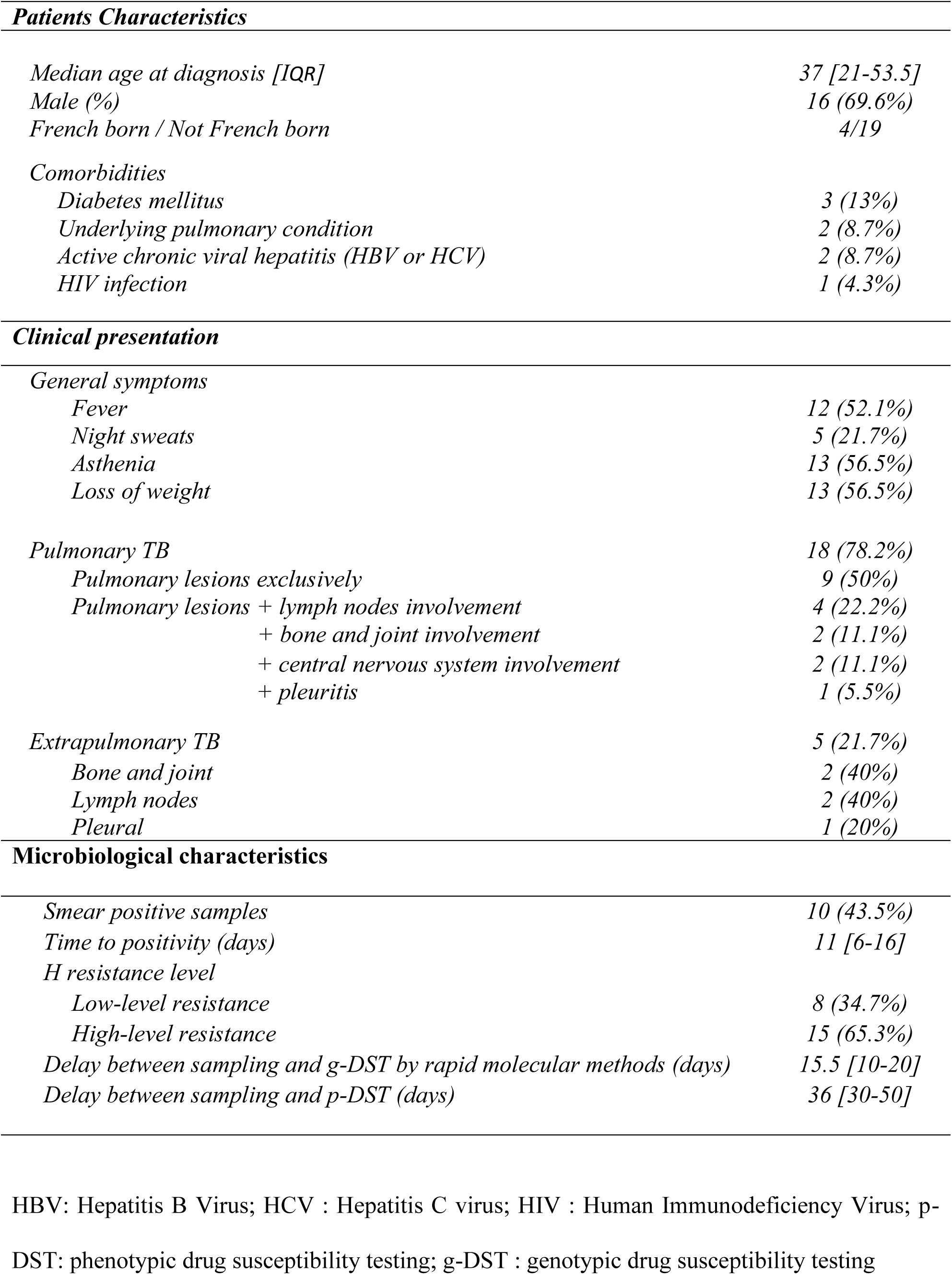
Main characteristics of the patients diagnosed with HR-TB.

The patients presented asthenia (n=13; 56.5%), weight loss (n=13; 56.5%), fever (n=12; 52.1%), and night sweats (n=5; 21.7%). Pulmonary TB occurred in 18 patients (78.3%) while 5 patients (21.7%) had extrapulmonary TB. Among the patients with pulmonary TB, 9 patients (50%) showed an isolated pulmonary involvement, 4 patients (22.2%) had an associated lymph nodes involvement, 2 patients (11.1%) had an associated vertebral involvement, 2 patients (11.1%) had a central nervous system involvement, and 1 patient (5.5%) had a pleural involvement. Among the 5 patients with extrapulmonary TB, 2 patients (40%) exhibited osteoarticular TB, 2 patients (40%) had lymph nodes TB and 1 patient (20%) had pleural TB. Among the 23 patients with HR-TB, 10 (43.5%) had smear positive samples, and the median [IQR] time to positivity of the culture was 11 days [6-16]. According to p-DST, 8 (34.7 %) patients had low-level H resistance and 15 (65.3 %) had high-level H resistance. The median [IQR] delay between sampling and g-DST performed on cultured Mtb was 15.5 days [10-20] and was significantly lower than the median [IQR] delay between sampling and p-DST of 36 days [30-50] (*p*<0.0001).

### Treatment and Outcome of HR-TB patients

Among the 23 patients with HR-TB, 16/23 (69.6%) received the standard regimen RHZE upon treatment initiation (Table 3, detailed in Table S2). An early detection of HR-TB on highly positive sputum in 2 (8.7%) patients resulted in the initiation of a FQ containing, H-free regimen from the start. Toxicity concerns were responsible for the start of non-conventional regimen for other patients. Following the molecular detection of H resistance in all patients, H doses were increased in 3 (13%) patients and H was switched to FQ in 10 (43.4%) patients. Additional phenotypic detection of H resistance resulted in 2 more switch from H to FQ. In total, 17 (73.9%) HR-TB patients received a FQ-containing regimen (13 received LFX, 3 received moxifloxacin (MOX) and 1 received both sequentially, Table S2). Toxicity or adverse events related to treatment modifications occurred in 5 (21.7%) patients, either due to hepatic cytolysis (n=2), epileptic seizure (n=1), tendinopathy (n=1), or neutropenia (n=1). During the continuation phase, 13 (56.5%) patients underwent treatment simplification combining two drugs, including R associated with either a FQ (n=8; 35%), Z (n=2; 8.7%), H (n=2; 8.7%) or E (n=1; 4.3%). The median (min-max) duration of these simplified regimens (from treatment modification to the end of anti-TB therapy) was 6 months (2-20). Overall, the median [IQR] treatment duration was 9 [6-12] months including 6 (26.1%) patients treated for 6 months, 8

**Table 3.**
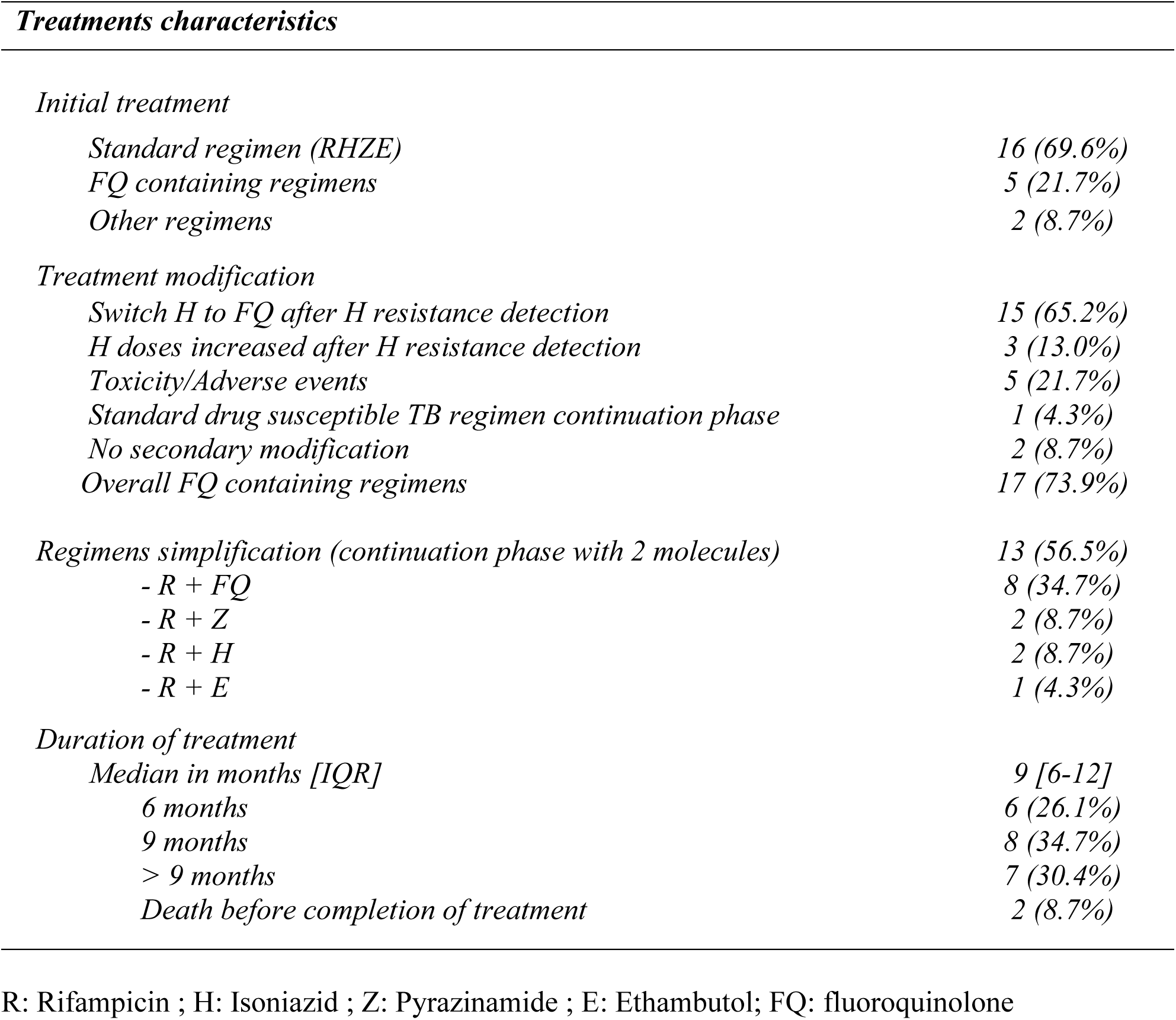
Main characteristics of the HR-TB patients’ treatment regimens.

| *Treatments characteristics* |  |
| --- | --- |
| *Initial treatment* |  |
| Standard regimen (RHZE) | 16 (69.6%) |
| FQ containing regimens | 5 (21.7%) |
| Other regimens | 2 (8.7%) |
| *Treatment modification* |  |
| Switch H to FQ after H resistance detection | 15 (65.2%) |
| H doses increased after H resistance detection | 3 (13.0%) |
| Toxicity/Adverse events | 5 (21.7%) |
| Standard drug susceptible TB regimen continuation phase | 1 (4.3%) |
| No secondary modification | 2 (8.7%) |
| Overall FQ containing regimens | 17 (73.9%) |
| *Regimens simplification (continuation phase with 2 molecules)* |  |
| - R + FQ | 8 (34.7%) |
| - R + Z | 2 (8.7%) |
| - R + H | 2 (8.7%) |
| - R + E | 1 (4.3%) |
| *Duration of treatment* |  |
| Median in months [IQR] | 9 [6-12] |
| 6 months | 6 (26.1%) |
| 9 months | 8 (34.7%) |
| > 9 months | 7 (30.4%) |
| Death before completion of treatment | 2 (8.7%) |

R: Rifampicin ; H: Isoniazid ; Z: Pyrazinamide ; E: Ethambutol; FQ: fluoroquinolone

(34.7%) patients treated for 9 months, and 7 (30.4%) treated > 9 months (range10 to 22). In 8 patients, a longer therapy was explained by the extrapulmonary localisation and/or by risk factors of treatment failure (extensive cavitary disease and/or slow conversion of smear/culture). However, the 7 patients who were treated for > 9 months had no associated risk factors for relapse or treatment failure.

Among the 23 patients, the outcome was favourable in 21 (91.3%) patients, while 2 (8.7%) patients died from TB unrelated cause before treatment completion, after 2 days and 21 days respectively (Table S2).

### Impact of microbiological results on HR-TB treatment

The anti-TB treatment was mostly initiated before the obtention of both p-DST and g-DST results; the median [IQR] delay between treatment initiation and p-DST was 32 days [28-45 days] and the median [IQR] delay between treatment initiation and g-DST was 10.5 days [4-12 days]. In 2 patients the g-DST was reported before treatment initiation, thus allowing an appropriate HR-TB management from the start (Figure 1A). Among the 23 patients with HR-TB, 19 (82.6%) were accurately detected using g-DST by rapid molecular methods. A total of 4 high-level H resistant cases were missed; 2 cases were presumed to have low-level H resistant, while 2 were presumed H susceptible (82.6% concordance between g-DST and p-DST).

**Figure 1:**
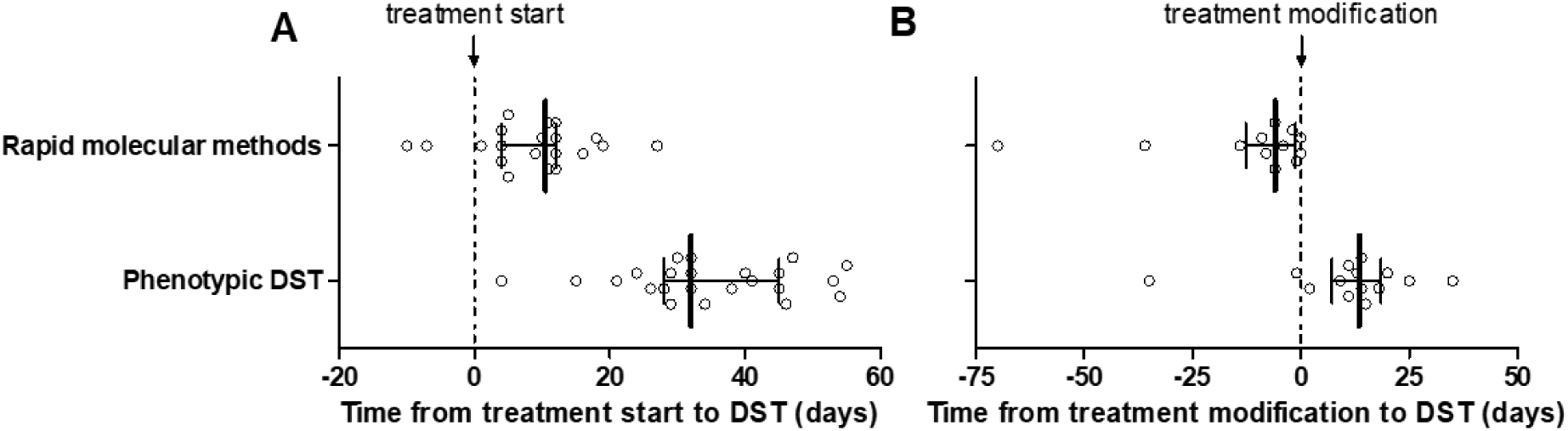
Delay between anti-TB treatment and drug susceptibility testing results. Genotypic DST by rapid molecular methods were performed on cultured Mtb, using the line probe assay GenoType MTBDRplus v2.0 test from November 2016 to August 2020 and the MDR/MTB ELITe MGB® Kit on the ELITe InGenius® platform from September 2020 to July 2022. Anti-TB treatment switch: replacement of H by FQ or increase in the dose of H.

The g-DST results generally led to the modification of anti-TB treatment, by replacing H by FQ or increasing the doses of H; the median [IQR] delay between g-DST results and treatment modification was 6 days [1-13 days]. In 2 patients, the treatment was only modified after p-DST results (Figure 1B); in 1 patient, the g-DST by rapid molecular method did not detect any H resistance associated mutation (*katG* deletion of amino acid 975, Table 1). In the other patient, for whom only the presumed low-level H resistance C-15T mutation on the *inhA* promoter was detected, the pursuit of the standard regiment combined with pharmacological monitoring was decided. In the other 2 patients, where H resistance mutations were not accurately detected by rapid molecular methods, 1 died before g-DST results (*katG* Q88P mutation), and 1, for whom g-DST only detected a mutation associated with low-level H resistance, LFX was implemented and then continued when p-DST revealed high-level H resistance (supported by NGS-based DST showing a double mutation of *inh*A locus: *inh*A C1673425T -15 + *inh*A T1674481G I194T).

## Discussion

The present study showed that rapid resistance detection tests allowed to accelerate the treatment adaptation in HR-TB patients. H resistance remains the most common type of resistance to anti-TB drugs. Global data on H resistance without concurrent R resistance were 7.4% in new TB cases and 11.4% in previously treated TB cases ^2^, whilst this series included 29 TB patients with HR-TB representing 4.4% of TB managed cases. Until recently, HR-TB was thought to have similar treatment outcomes compared to drug-susceptible TB. However, many studies support that HR-TB is associated with higher rates of treatment failure and relapse than drug-susceptible TB ^18^. The increasing risk of MDR-TB in case of relapse, urged the elaboration of guidelines for appropriate management of HR-TB ^3^. To fasten the treatment adaptation, rapid testing for H resistance, in addition to R resistance, is largely encouraged by the WHO ^4^. The present results thus confirm previous reports showing that g-DST compared to p-DST significantly accelerates HR-TB diagnosis and appropriate anti-TB implementation, and should therefore be performed on routine basis by all TB diagnosis laboratories.

Though g-DST by rapid molecular methods may speed results by several weeks, the lack of sensitivity of H resistance remains a challenge ^12^.This could be explained by the fact that clinical diagnosis companies mainly focus on R resistance detection as (i) MDR-TB always include resistance to R; (ii) the genetic support of R resistance is restricted to a short nucleotide sequence of *rpo*B locus; (iii) as a corollary there is an excellent feasibility of molecular tests ^4,19^. Herein, the performances of rapid molecular R resistance detection were excellent. Conversely, regarding H resistance, the majority of rapid molecular methods only target codon 315 of *katG* locus (associated with high-level H resistance), leaving undetected other polymorphisms, frameshifts, and deletions on *katG,* though probably associated with phenotypic resistance ^5^. This is linked to the genetic basis of H resistance; the heterogeneous performance of currently developed PCR-based assays for H-resistance detection can be explained by multi-locus spread genetic modifications ^6,13^. This issue seems to have been overpassed by the NGS-based assay Deeplex Myc-TB®, which herein displayed performances very similar to p-DST. Nevertheless, this assay needs every-day access to NGS services to be able to provide results in a time frame similar to rapid molecular tests.

Previously very useful to detect the different levels of H-resistance, p-DST was essential to determine HR-TB treatment regimens; using high-dose of H in patients having low-level H-resistance, or replacing H by LFX in patients having high-level H-resistance. In order to prevent inappropriate HR-TB management in case of misdiagnosis of H high-level resistance, the WHO pragmatically recommends the replacement of H by LFX within the anti-TB regimen, regardless of the H resistance level ^3^. In the present series, 17/23 patients received FQ containing regimen in accordance with international guidelines. Since LFX containing regimen are increasingly implemented, it is more important to perform an accurate detection of H resistance than evaluating the level of H resistances. Although FQ resistance is still very low, 3/654 herein, to ensure safe implementation of FQ treatment, there is a need for a rapid detection of FQ resistance, in addition to a rapid and accurate resistance detection of the first-line anti-TB drugs. From this respect, to address the need of combined detection on multiple resistance markers, exhaustive NGS-based methods such as Deeplex® Myc-TB or WGS reveal to be more useful than prior developed real-time PCR-based methods. Herein, the excellent performances of NGS-based methods were confirmed for anti-TB drugs resistance detection, consistent with the target product profile requested for Mtb DST ^20^, and thus providing a culture-free DST solution for TB patients.

In this study, the median duration of anti-TB therapy was higher than the one recommended by the WHO 2018 guidelines: a duration of 6 months for LFX containing regimen ^21^. However, previous guidelines (2014) recommended a treatment duration of 6 to 9 months ^22^, thus, the prolongation to 9 months of treatment in absence of associated risk factors observed in our patients might be explained by the heterogeneous comprehension of successive guidelines. Unexpectedly, a regimen reduction to bitherapy was observed in 11 patients not managed with the RHZE standard regimen, including 8 patients with R+FQ regimen. This therapeutic option may be explained by the misinterpretation of the WHO guidelines, tending to mimic the switch to bitherapy performed upon the standard regimen, as well as by the intolerance or fear of intolerance in 6 months multi-antibiotic treatment. However, the favourable outcome observed for all the patients concerned by this simplification, may suggest that FQ containing regimens could be simplified upon the continuation phase.

Our study has limitations, firstly the enrolling centres were close geographically and linked to a single laboratory diagnosis, thus, it did not entirely reflect the French TB diagnosis and management landscape. Secondarily, during the 5 years of the enrolling period, changes of the diagnosis methods were performed as well as changes of TB management guidelines, probably explaining the heterogeneity of treatments. However, our results were in line with a previous French multi-centre assessment covering the first part of our enrolling period ^11^, revealing, in our case, a better implementation of g-DST and also a better compliance with the international HR-TB management guidelines, consistent with the 2018 WHO new recommendations.

The present study highlighted the importance of g-DST to safely engage HR-TB patients on adapted treatment. Also, NGS-based g-DST showed almost perfect concordance with p-DST, thus providing rapid and safe culture-free DST alternative. Moreover, this study revealed an important heterogeneity regarding patient management, probably due to the clinical complexity of patients, but also to misinterpretation of recent guidelines. We identified a need of simplified regimens to prevent toxicity and improve treatment adherence, in the context of TB management secured by accurate NGS-based resistance detection tools.

## Supporting information

Figure S1: Microbiological description of the TB patient cohort.

## Data Availability

All data produced in the present study are available upon reasonable request to the authors

## Acknowledgements

We thank Shanez Haouari (Direction de la Recherche en Santé, Hospices Civils de Lyon) for help in manuscript preparation.

---

**Figure S1:**
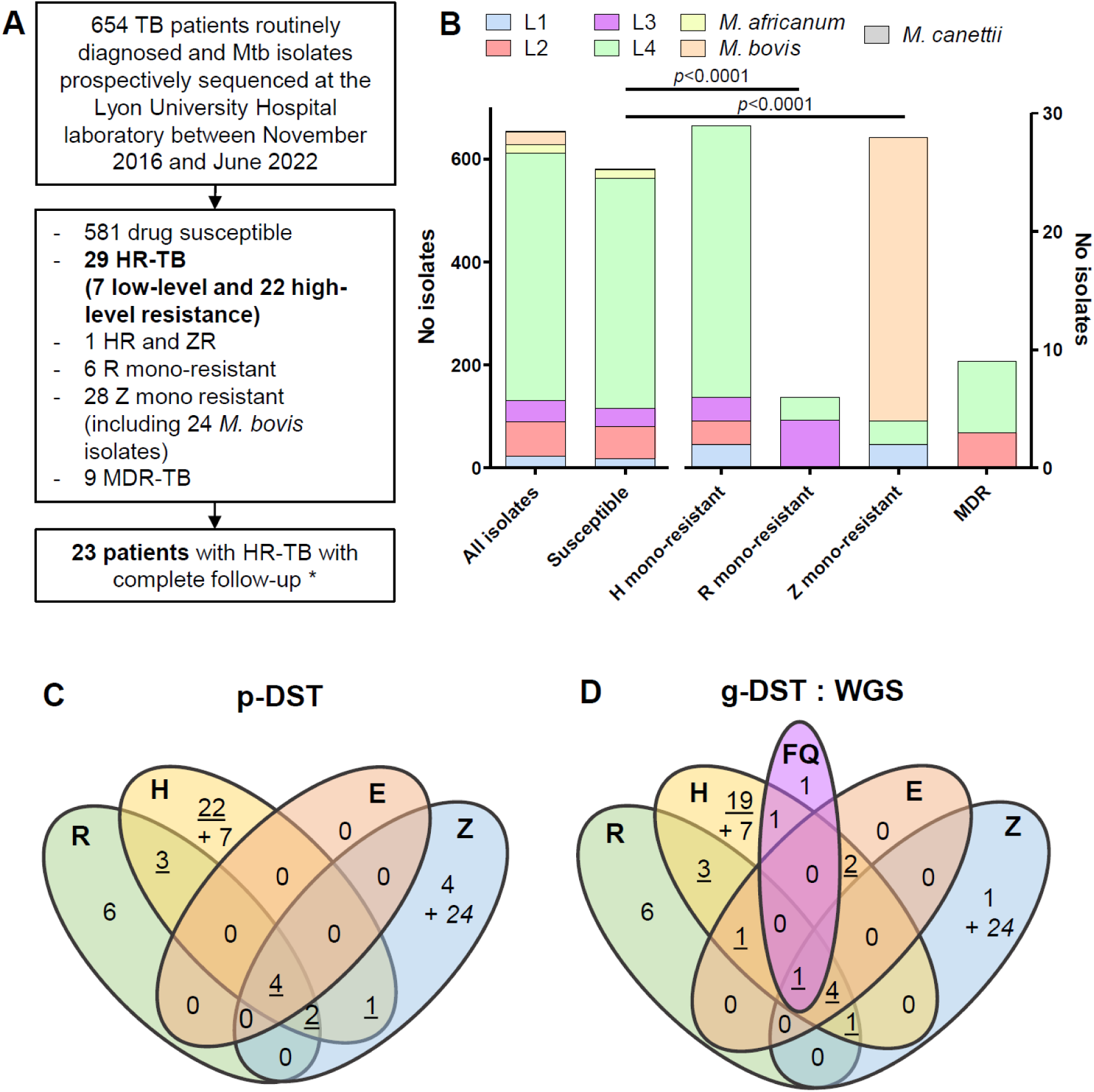
Microbiological description of the TB patient cohort. A. Resistance profile of TB patients diagnosed at the Lyon University Hospital. *6 patients were followed-up until the end of treatment in a different center. B. Distribution of Mtb lineages according to the resistance profile. C and D. Distribution of the 73 Mtb isolates resistant to at least one first-line drug by phenotypic drug susceptibility testing (p-DST, C) or genotypic DST (g-DST) performed by whole genome sequencing (WGS). Detection of resistance to fluoroquinolone was also performed by g-DST using WGS. R, rifampicin; H, isoniazid; Z, pyrazinamide; E, ethambutol; FQ, fluoroquinolone. Underlined: H high-level resistance; italics: *M. bovis* isolates.

**Table S2.**
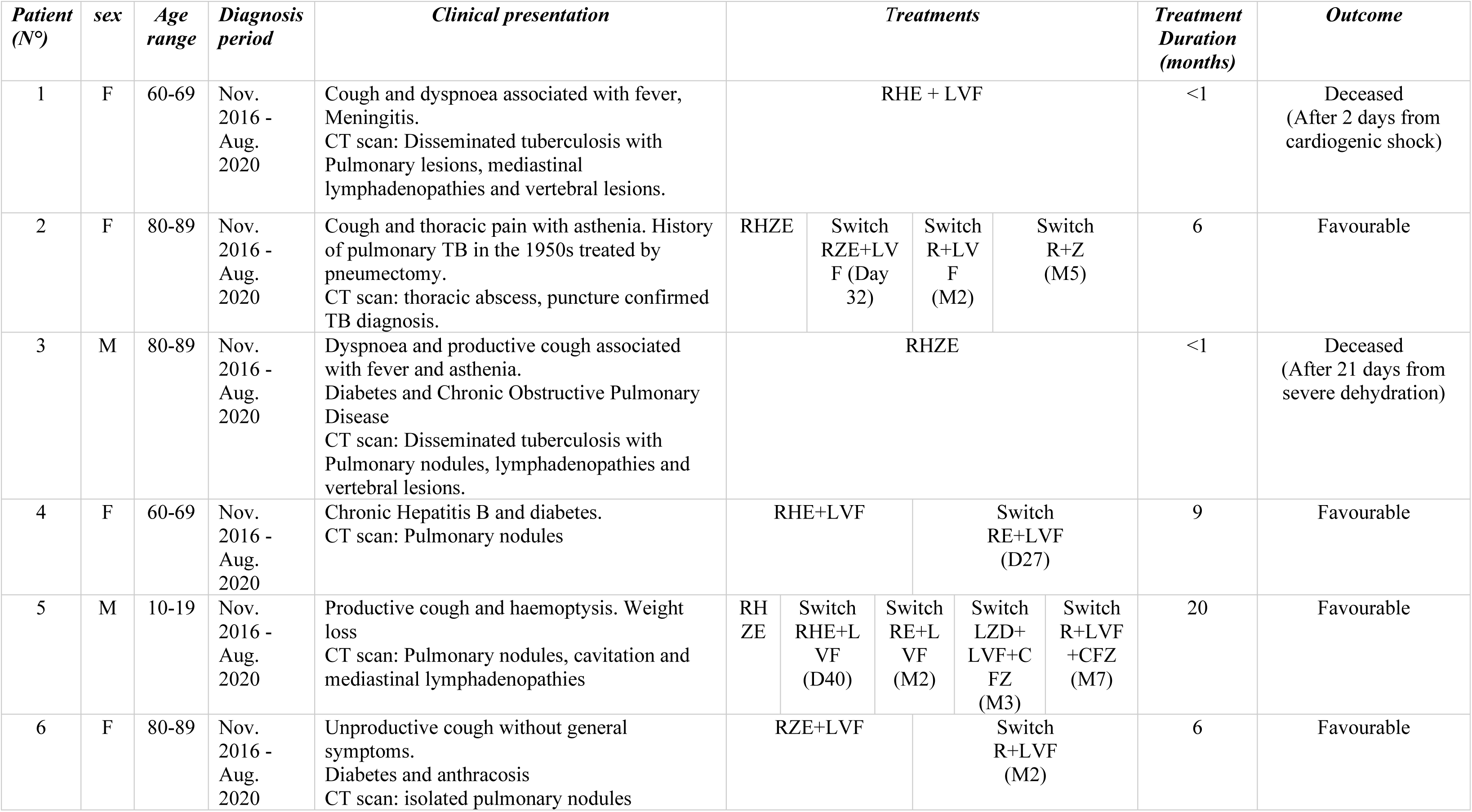
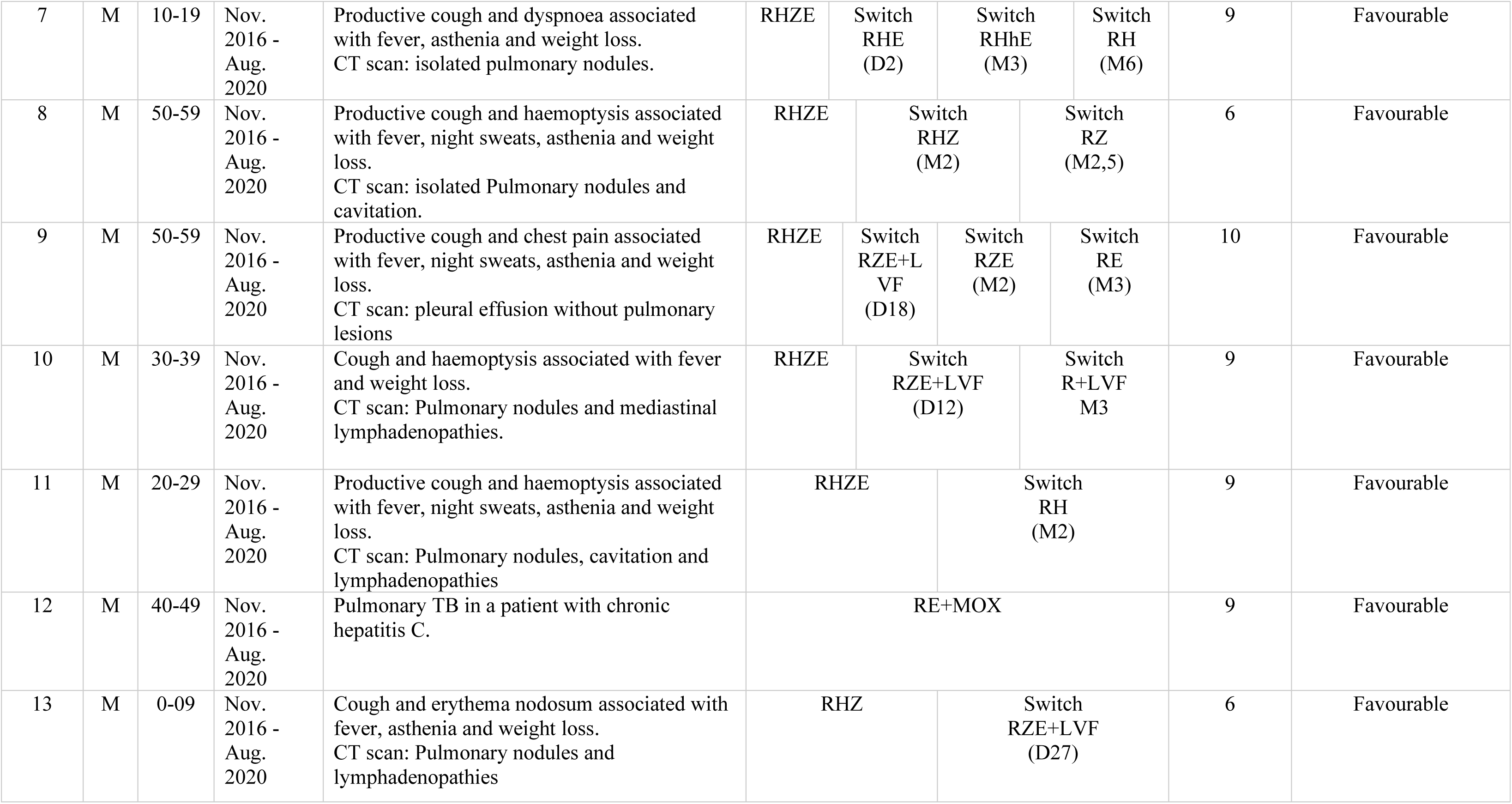
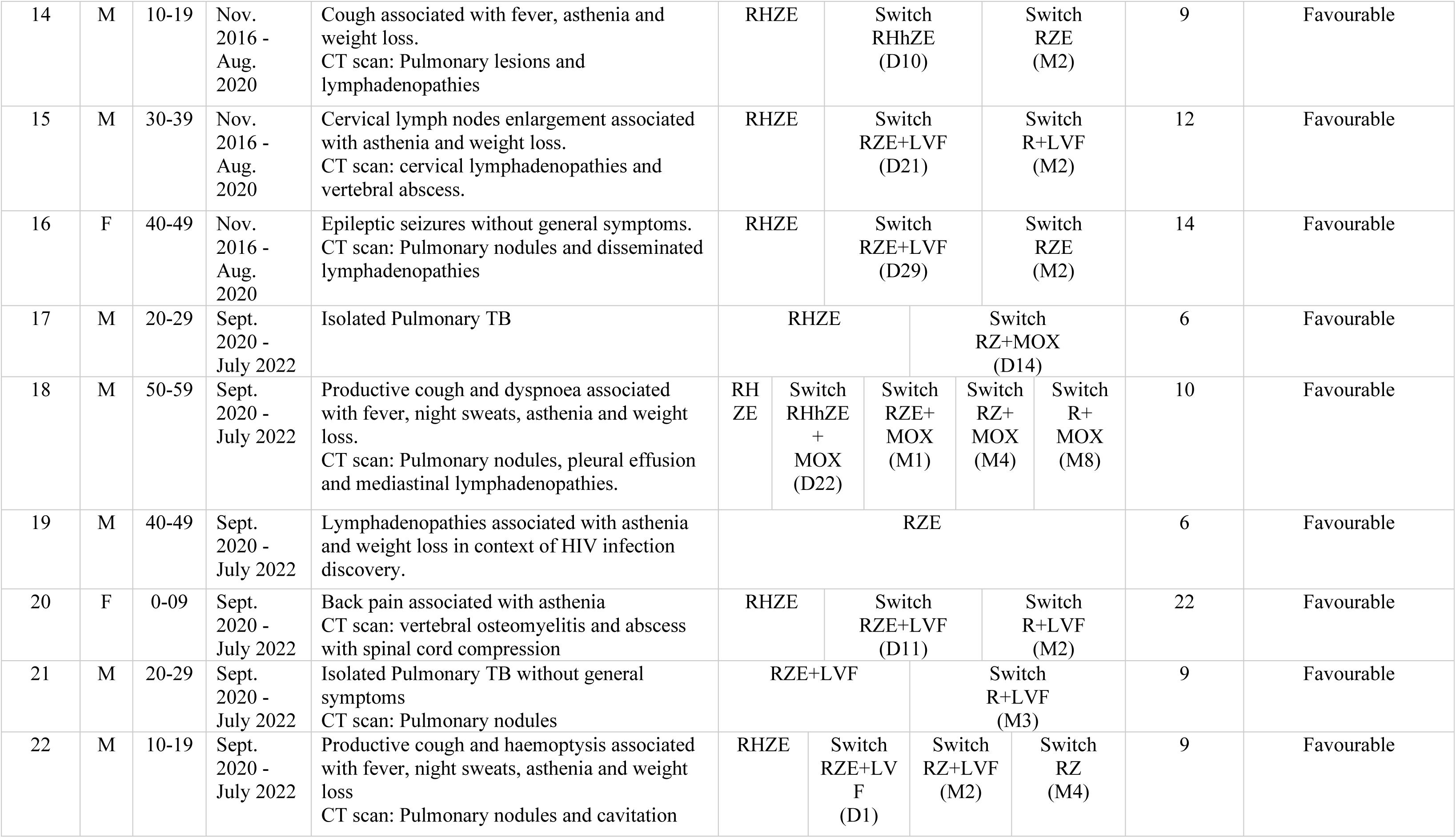
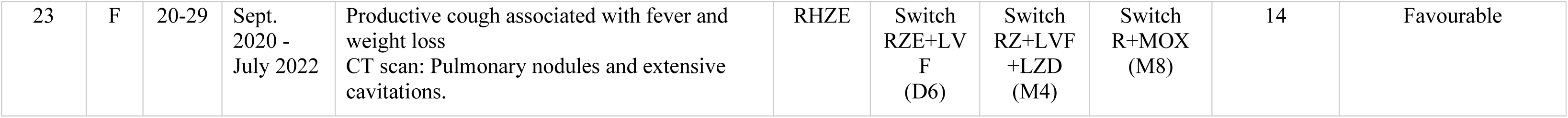
Patients detailed clinical presentation and therapeutic regimens.

## References

1 World Health Organization, Geneva. Global tuberculosis report 2022. Available at https://www.who.int/publications-detail-redirect/9789240061729. Accessed December 13, 2022, 2022.

2 Dean Anna S., Zignol Matteo, Cabibbe Andrea Maurizio, Falzon Dennis, Glaziou Philippe, Cirillo Daniela Maria, et al. Prevalence and genetic profiles of isoniazid resistance in tuberculosis patients: A multicountry analysis of cross-sectional data. PLOS Med 2020;17(1):e1003008. Doi: 10.1371/journal.pmed.1003008.

3. World Health Organization, Geneva. WHO consolidated guidelines on drug-resistant tuberculosis treatment. Available at https://apps.who.int/iris/rest/bitstreams/1211676/retrieve. 2019.

4 World Health Organization, Geneva. WHO consolidated guidelines on tuberculosis. Module 3: diagnosis - rapid diagnostics for tuberculosis detection, 2021 update. Geneva, Switzerland; 2021.

5 Rivière E., Whitfield M. G., Nelen J., Heupink T. H., Van Rie A. Identifying isoniazid resistance markers to guide inclusion of high-dose isoniazid in tuberculosis treatment regimens. Clin Microbiol Infect Off Publ Eur Soc Clin Microbiol Infect Dis 2020;26(10):1332–7. Doi: 10.1016/j.cmi.2020.07.004.

6 Vogensen Vanessa B., Anthony Richard M., Kerstjens Huib A.M., Tiberi Simon, De Steenwinkel Jurriaan E.M., Akkerman Onno W. The case for expanding worldwide access to point of care molecular drug susceptibility testing for isoniazid. Clin Microbiol Infect 2022;28(8):1047–9. Doi: 10.1016/j.cmi.2022.03.033.

7 Dooley Kelly E., Miyahara Sachiko, von Groote-Bidlingmaier Florian, Sun Xin, Hafner Richard, Rosenkranz Susan L., et al. Early Bactericidal Activity of Different Isoniazid Doses for Drug-Resistant Tuberculosis (INHindsight): A Randomized, Open-Label Clinical Trial. Am J Respir Crit Care Med 2020;201(11):1416–24. Doi: 10.1164/rccm.201910-1960OC.

8 Jouet Agathe, Gaudin Cyril, Badalato Nelly, Allix-Béguec Caroline, Duthoy Stéphanie, Ferré Alice, et al. Deep amplicon sequencing for culture-free prediction of susceptibility or resistance to 13 anti-tuberculous drugs. Eur Respir J 2021;57(3):2002338. Doi: 10.1183/13993003.02338-2020.

9 Walker Timothy M., Miotto Paolo, Köser Claudio U., Fowler Philip W., Knaggs Jeff, Iqbal Zamin, et al. The 2021 WHO catalogue of *Mycobacterium tuberculosis* complex mutations associated with drug resistance: A genotypic analysis. Lancet Microbe 2022;3(4):e265–73. Doi: 10.1016/S2666-5247(21)00301-3.

10. World Health Organization Geneva. The use of next-generation sequencing technologies for the detection of mutations associated with drug resistance in Mycobacterium tuberculosis complex: technical guide. 2018.

11 Bachir Marwa, Guglielmetti Lorenzo, Tunesi Simone, Billard-Pomares Typhaine, Chiesi Sheila, Jaffré Jérémy, et al. Molecular detection of isoniazid monoresistance improves tuberculosis treatment: A retrospective cohort in France. J Infect 2022;85(1):24–30. Doi: 10.1016/j.jinf.2022.05.017.

12 Genestet Charlotte, Hodille Elisabeth, Berland Jean-Luc, Ginevra Christophe, Bryant Juliet E., Ader Florence, et al. Whole-genome sequencing in drug susceptibility testing of *Mycobacterium tuberculosis* in routine practice in Lyon, France. Int J Antimicrob Agents 2020;55(4):105912. Doi: 10.1016/j.ijantimicag.2020.105912.

13 Hodille Elisabeth, Genestet Charlotte, Delque Thomas, Ruffel Luna, Benito Yvonne, Fredenucci Isabelle, et al. The MTB/MDR ELITe MGB® Kit: Performance Assessment for Pulmonary, Extra-Pulmonary, and Resistant Tuberculosis Diagnosis, and Integration in the Laboratory Workflow of a French Center. Pathog Basel Switz 2021;10(2). Doi: 10.3390/pathogens10020176.

14 Hodille Elisabeth, Maisson Audey, Charlet Laurine, Bauduin Clyde, Genestet Charlotte, Fredenucci Isabelle, et al. Evaluation of Xpert MTB/RIF Ultra performance for pulmonary tuberculosis diagnosis on smear-negative respiratory samples in a French centre. Eur J Clin Microbiol Infect Dis Off Publ Eur Soc Clin Microbiol 2019;38(3):601–5. Doi: 10.1007/s10096-018-03463-1.

15 Feuerriegel Silke, Kohl Thomas A., Utpatel Christian, Andres Sönke, Maurer Florian P., Heyckendorf Jan, et al. Rapid genomic first- and second-line drug resistance prediction from clinical *Mycobacterium tuberculosis* specimens using Deeplex-MycTB. Eur Respir J 2021;57(1):2001796. Doi: 10.1183/13993003.01796-2020.

16 Genestet Charlotte, Hodille Elisabeth, Bernard Albin, Vallée Maxime, Lina Gérard, Le Meur Adrien, et al. Consistency of *Mycobacterium tuberculosis* Complex Spoligotyping between the Membrane-Based Method and In Silico Approach. Microbiol Spectr 2022;10(3):e0022322. Doi: 10.1128/spectrum.00223-22.

17 Bradley Phelim, Gordon N. Claire, Walker Timothy M., Dunn Laura, Heys Simon, Huang Bill, et al. Rapid antibiotic-resistance predictions from genome sequence data for *Staphylococcus aureus* and *Mycobacterium tuberculosis*. Nat Commun 2015;6(1):10063. Doi: 10.1038/ncomms10063.

18 Menzies Dick, Benedetti Andrea, Paydar Anita, Royce Sarah, Madhukar Pai, Burman William, et al. Standardized treatment of active tuberculosis in patients with previous treatment and/or with mono-resistance to isoniazid: a systematic review and meta-analysis. PLoS Med 2009;6(9):e1000150. Doi: 10.1371/journal.pmed.1000150.

19 Jagielski Tomasz, Bakuła Zofia, Brzostek Anna, Minias Alina, Stachowiak Radosław, Kalita Joanna, et al. Characterization of Mutations Conferring Resistance to Rifampin in *Mycobacterium tuberculosis* Clinical Strains. Antimicrob Agents Chemother 2018;62(10):e01093–18. Doi: 10.1128/AAC.01093-18.

20 MacLean Emily Lai-Ho, Miotto Paolo, González Angulo Lice, Chiacchiaretta Matteo, Walker Timothy M., Casenghi Martina, et al. Updating the WHO target product profile for next-generation *Mycobacterium tuberculosis* drug susceptibility testing at peripheral centres. PLOS Glob Public Health 2023;3(3):e0001754. Doi: 10.1371/journal.pgph.0001754.

21 World Health Organization, Geneva. WHO treatment guidelines for isoniazid-resistant tuberculosis: supplement to the WHO treatment guidelines for drug-resistant tuberculosis. 2018.

22 World Health Organization, Geneva. Companion handbook to the WHO guidelines for the programmatic management of drug resistant tuberculosis 2014.

