## Supplementary figures and images for "Rapid resistance detection is reliable for prompt adaptation of isoniazid resistant tuberculosis management"

### Figure S1: Microbiological description of the TB patient cohort.

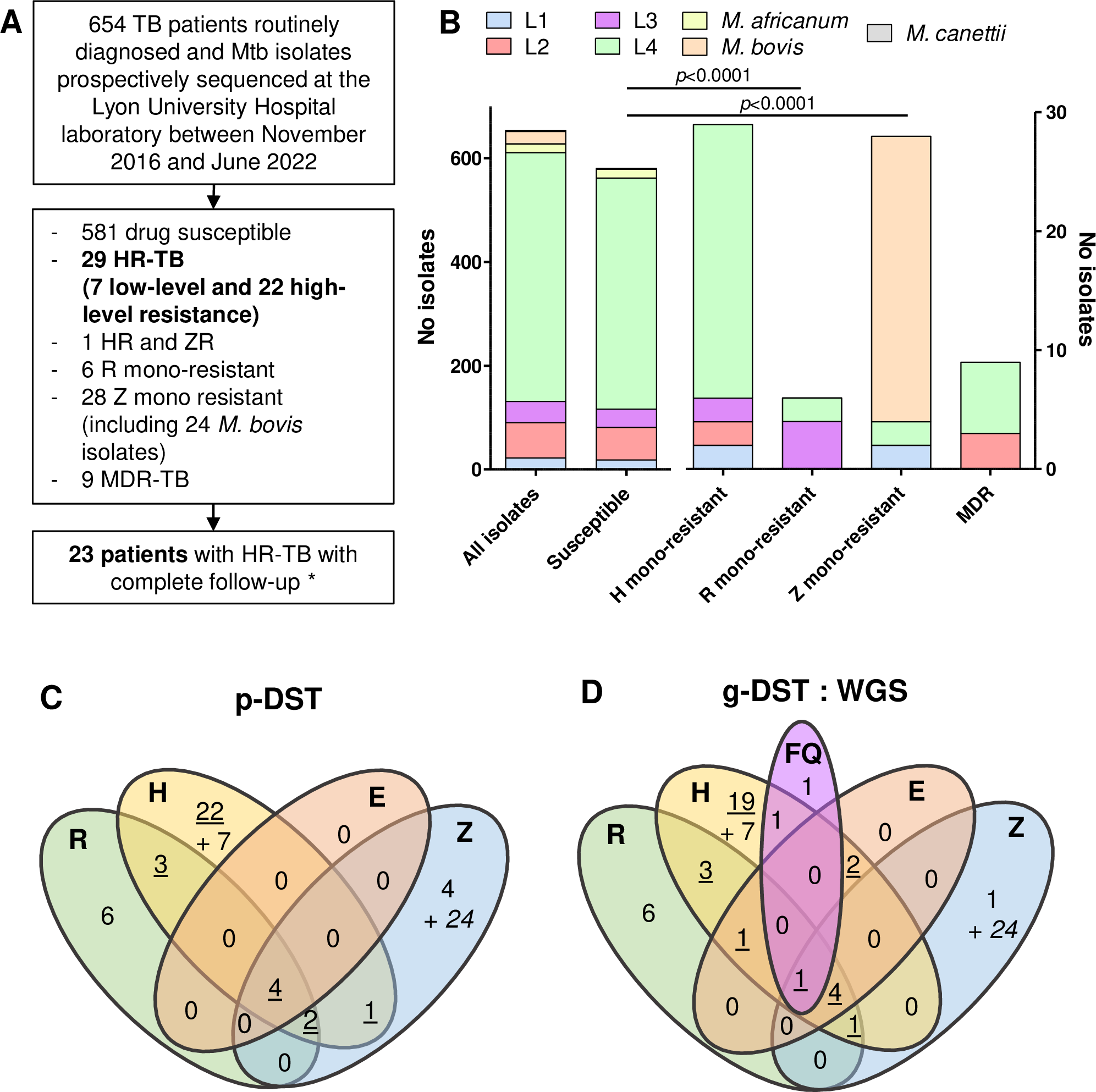
